# Performance of family history-based colorectal cancer screening criteria by race and age at diagnosis in the Disparities and Cancer Epidemiology (DANCE) study

**DOI:** 10.64898/2026.06.16.26355827

**Authors:** Kristen S. Purrington, Cydnie Martin, Angela S. Wenzlaff, Julie J. Ruterbusch, Siddhi Patil, Stephanie S. Pandolfi, Ivan Samayoa, Ann G. Schwartz, Mei-Chin Hsieh, Elena M. Stoffel, Laura Rozek

## Abstract

**Importance:** Family history (FH) and age are the primary criteria employed for early colorectal cancer (CRC) risk stratification. We evaluated how well these criteria identify individuals diagnosed with CRC across age and racial groups.

**Objective:** To evaluate the performance of FH- and age-based screening criteria for identifying individuals with CRC, with attention to differences by race and age at diagnosis.

**Design, Setting, and Participants:** This case-control and case-only analysis used data from the Disparities and Cancer Epidemiology (DANCE) cohort, a population-based study of invasive CRC cases diagnosed from 2013 to 2022, recruited through the Metropolitan Detroit Cancer Surveillance System and the Louisiana Tumor Registry. Analyses included 1,158 non-Hispanic Black (NHB) and non-Hispanic White (NHW) CRC cases and 1,434 cancer-free controls from the Inflammation Health and Lung Epidemiology (INHALE) study, enrolled from the same Detroit catchment area. Data were analyzed in 2025.

**Exposures:** Self-reported cancer FH among first-degree (FD) relatives and grandparents, summarized into three FH-based screening criteria: at least one FD relative with CRC (colon early-screening criterion), any FH of Lynch syndrome-related cancers, and meeting NCCN criteria for Lynch syndrome genetic testing.

**Main Outcomes and Measures:** Proportion of cases meeting each FH-based screening criterion stratified by race and age at diagnosis (<45, 45-49, 50-64, ≥65 years); case-only odds ratios for younger age at diagnosis; and case-control odds ratios for CRC associated with each criterion, with race-by-age interaction tested.

**Results:** Cancer FH burden differed by age at diagnosis across both racial groups. First degree (FD) CRC FH was highest among NHB CRC cases diagnosed before age 45 (22.6%) and lowest in those diagnosed at ages 45–49 (8.2%), while NHW participants reported more CRC FH with older age at diagnosis (p-interaction=0.011). In case-control analyses, having at least one FD relative with CRC was associated with higher odds of CRC before age 45 among NHB (OR=1.44, 95% CI 1.09–1.89) but not NHW individuals. The proportion of cases diagnosed before age 45 with a FD CRC FH was low, though markedly higher in NHB than NHW individuals (22.6% vs. 4.0%). While the proportion was slightly higher when including FH of any Lynch syndrome-related cancers (NHB: 24.5%, NHW: 10.0%), the proportion of controls with a FD FH of these cancers also increased.

**Conclusions and Relevance:** Current family history-based criteria fail to identify the majority of individuals diagnosed with CRC before age 45, with performance varying substantially by race, highlighting the urgent need for more equitable and effective approaches to early-onset CRC risk stratification.

**Key Points:** 

**Question:** How well do family history (FH)-based criteria identify individuals diagnosed with colorectal cancer (CRC), and does performance differ by race and age at diagnosis?

**Findings:** In this case-control study of 1,158 colorectal cancer cases and 1,434 cancer-free controls, current FH-based criteria failed to identify the majority of cases diagnosed before age 45, capturing fewer than one-quarter of non-Hispanic Black and fewer than 5% of non-Hispanic White early-onset cases. A statistically significant race-by-age interaction was observed.

**Meaning:** Family history-based criteria perform poorly for identifying early-onset CRC and differ by race, underscoring the need for more equitable and effective screening strategies.

## Introduction

Incidence of early-onset colorectal cancer (CRC) is increasing in the US, with rates in adults aged 20-49 years rising by 3% annually from 2013-2022, and CRC is now the leading cause of cancer death in this age group^1^. Young patients with CRC are more likely to present with stage III or IV disease than older patients^3^, reflecting diagnostic delays when early-onset cases go unrecognized. Family history (FH)- and age-based criteria are the principal mechanism for identifying younger individuals as candidates for early screening, yet most individuals diagnosed with CRC report no family history of the disease, and whether current criteria adequately capture FH burden at younger ages has not been systematically evaluated in population-based data.

Having one or more first-degree (FD) relatives with CRC doubles lifetime risk, and risk is substantially higher when affected relatives were diagnosed at younger ages^4^. Hereditary CRC syndromes such as Lynch syndrome increase CRC risk approximately 10-fold; National Comprehensive Cancer Network (NCCN) guidelines recommend that individuals from Lynch syndrome families begin colonoscopy screening between ages 20 and 25, or 2-5 years prior to the earliest cancer diagnosis in the family if it occurred before age 25^5^. Together, FD relative history, Lynch-related cancers in the family, and NCCN high-risk criteria represent the primary tools clinicians use to recommend earlier screening initiation outside the average-risk population.

Modeling studies suggest that systematic application of FH-based CRC risk stratification could reduce cancer morbidity and mortality, but empirical data supporting these projections are limited^6^. A fundamental challenge is that FH criteria depend on patients’ knowledge of their family cancer history, which is often incomplete: studies have found sensitivity for identifying CRC in FD relatives as low as 0.57^7^. Familial risk profiles, Lynch syndrome risk models, and universal tumor screening can help identify high-risk patients, but many cases are still missed due to patient reluctance, reliance on physician prompting, and practical barriers to comprehensive FH collection^8^.

We used the Disparities and Cancer Epidemiology (DANCE) population-based cohort of CRC patients to examine the prevalence of cancer FH patterns used to identify candidates for early screening. We evaluated how well these criteria perform across categories of age at diagnosis and race. We identified gaps in current CRC risk stratification approaches that may contribute to under-detection of individuals at increased risk for developing CRC and highlight the need for more effective strategies for earlier screening initiation.

## Methods

### Design, Setting, and Participants

DANCE is a population-based cohort of non-Hispanic Black (NHB) and non-Hispanic White (NHW) invasive CRC. Cases were recruited through the Metropolitan Detroit Cancer Surveillance System (MDCSS) and the Louisiana Tumor Registry (LTR) cancer registries. The MDCSS catchment area covers Wayne, Oakland, and Macomb counties in southeast Michigan, and the LTR catchment area covers the entire state of Louisiana. Patients were eligible if they were diagnosed with invasive CRC from 2013-2022, lived within the MDCSS or LTR catchment areas, and self-identified as NHB or NHW. All DANCE participants completed baseline questionnaires either online, over the phone with a trained interviewer, or by returning a written survey. Some DANCE cases were originally enrolled in the Detroit Research on Cancer Survivors study (n=528), an MDCSS-based cohort of Black cancer survivors^9,10^. Participants provided informed consent, and study protocols were approved by Institutional Review Boards at the University of Michigan, Georgetown University, Wayne State University, and Louisiana State University.

### INHALE controls

The INHALE study has been described in detail previously^11^. Volunteer controls included in this analysis were NHB or NHW, aged 21–89 years, and enrolled from the MDCSS catchment area. INHALE study criteria required that controls had never taken amiodarone or been diagnosed with bronchiectasis or cystic fibrosis, carried health insurance, never had a surgical lung resection, and were never diagnosed with lung cancer. We further excluded all controls reporting a personal history of any cancer type (n=249).

### Patient Data Collection

For DANCE, cancer FH was collected in the baseline questionnaire, including information on the number of FD (FD) relatives, whether any FD relatives or grandparents had ever been diagnosed with cancer, and family members’ ages at cancer diagnosis. Participants completing the online survey were asked about FH of breast, bladder, cervix, colorectal, esophageal, endometrial, head and neck, kidney, leukemia, liver, lung, lymphoma, melanoma, myeloma, ovarian, pancreatic, stomach, testis, and thyroid. Participants also had the option to indicate that the cancer site was unknown or to write in another cancer type. Write-in responses for cancer type were coded by 2 independent reviewers and any discordance was reviewed with the study PI to determine the final category. Data on participants’ age at diagnosis, sex, and education were obtained from baseline questionnaires and data linkage with MDCSS and LTR. For INHALE controls, questionnaires collected data on cancer FH among first degree relatives, participant sex and age at enrollment. Race and ethnicity were self-reported by participants at study enrollment using fixed categories from the baseline questionnaire (non-Hispanic Black, non-Hispanic White, Hispanic, other, or prefer not to answer). Self-reported race and ethnicity were collected because of well-documented racial disparities in CRC incidence, survival, and screening uptake that are central to the study aims.

### Family history definitions

Cancer types were grouped into etiologically related categories including colorectal, other gastrointestinal (GI: esophageal, stomach, pancreatic), estrogen-driven (breast, ovarian), prostate, hematologic (leukemia, lymphoma, myeloma), environmentally-driven by agents such as smoking, viral infection, or radiation (lung, kidney, head and neck, liver, melanoma, cervix, thyroid), other rare cancers (bone, brain, eye, soft tissue, testis), or unknown cancer type. Lynch syndrome-related cancers were defined as colorectal, endometrial, ovarian, stomach or pancreatic cancers. Based on clinical guidelines, we considered having at least one FD relative with CRC as an indication for initiating CRC screening before age 45. NCCN criteria (version 4.2024) were used to define FH-based eligibility criteria for genetic testing. We did not include personal cancer history criteria in an effort to capture information that would have been used to direct CRC screening recommendations before participants were diagnosed with CRC. This included at least two close relatives with a Lynch-related cancer at any age or at least one close relative diagnosed under age 50. We defined close relatives using FD relatives alone and also using FD relatives plus grandparents, resulting in two versions of eligibility criteria.

### Statistical analysis

All analyses were conducted in R v4.5.2 (https://cran.r-project.org). Distributions of demographic and FH variables were calculated as frequencies and percentages for categorical variables and as medians and ranges for continuous variables. Family histories were summarized according to the proportion of participants reporting any relatives with a given cancer type/category, and results did not change appreciably if we instead considered number of relatives. Differences by age group at diagnosis were tested with chi-square or Fisher exact tests as appropriate. Case-only association analyses between FH-based screening categories and age group at onset were performed using unconditional logistic regression. Statistical interactions between race and age group at diagnosis were tested by incorporating interaction terms into the model. Case-control analyses were conducted using logistic regression. P-values ess than 0.05 were considered statistically significant, between 0.05 and 0.10 were considered marginally significant, and P values > 0.10 are referred to in the text as not significant (n.s.).

## Results

A total of 1,158 Disparities and Colorectal Cancer Epidemiology (DANCE) study participants with a personal history of CRC were included in the analysis (**Table 1**). The majority of cases were in the 50-64 (47.6%) and 65+ (34.6%) years at diagnosis age groups, although ∼9% of cases belonged to each of the <45 or 45-49 years at diagnosis age groups. Most DANCE participants were NHW (56.4%), resided in Michigan (78.5%), were male (50.6%), and had at least some college education (66.7%), with no significant differences by age at diagnosis. However, while not significant, there were more NHB participants in the <45 and 65+ age groups (47.6% NHB) than in the 45-49 and 50-64 age groups (40.5% NHB). The median number of reported FD relatives affected with cancer increased significantly as age group increased (P <0.001).

**Table 1.** Demographic characteristics of DANCE participants.

| Characteristic | <45<br>n (%) | 45-49<br>n (%) | 50-64<br>n (%) | 65+<br>n (%) | p-val |
| --- | --- | --- | --- | --- | --- |
| Total | 103 (100) | 103 (100) | 551 (100) | 401 (100) |  |
| Race |  |  |  |  |  |
| Non-Hispanic Black | 50 (48.5) | 42 (40.8) | 223 (40.5) | 190 (47.4) | 0.12 |
| Non-Hispanic White | 53 (51.5) | 61 (59.2) | 328 (59.5) | 211 (52.6) |  |
| State of residence |  |  |  |  |  |
| Michigan | 83 (80.6) | 80 (77.7) | 436 (79.1) | 310 (77.3) | 0.86 |
| Louisiana | 20 (19.4) | 23 (22.3) | 115 (20.9) | 91 (22.7) |  |
| Sex |  |  |  |  |  |
| Female | 55 (53.4) | 50 (48.5) | 263 (47.7) | 204 (50.9) | 0.65 |
| Male | 48 (46.6) | 53 (51.5) | 288 (52.3) | 197 (49.1) |  |
| Highest level of education |  |  |  |  |  |
| Less than HS | 5 (4.9) | 6 (5.8) | 33 (6.0) | 36 (9.0) | 0.51 |
| HS | 21 (20.4) | 22 (21.4) | 150 (27.2) | 96 (23.9) |  |
| Some college | 58 (56.3) | 60 (58.2) | 274 (49.7) | 198 (49.4) |  |
| Professional/graduate | 18 (17.5) | 15 (14.6) | 84 (15.2) | 65 (16.2) |  |
| Unknown | 1 (1.0) | 0 (0) | 10 (1.8) | 6 (1.5) |  |
| No. of first degree relatives |  |  |  |  |  |
| Median (Min-Max) | 4 (2-17) | 5 (2-16) | 6 (2-21) | 7 (2-24) | <0.001 |

We first evaluated differences in FH patterns by race in the DANCE study (**Table 2**). Considering only FD relatives or FD relatives and grandparents combined (FD+GP), the most commonly reported cancers were estrogen-driven (FD: 19.8%, FD+GP: 24.6%) and environmentally-driven (FD: 20.8%, FD+GP: 27.8%) cancers, followed by colorectal (FD: 13.6%, FD+GP: 19.3%), prostate (FD: 11.5%, FD+GP: 14.2%), and unknown cancer types (FD: 14.8%, FD+GP: 20.6%). The proportion of cases reporting FH among FD relatives of estrogen-driven (22.8% vs 17.5%, P = 0.025), hematologic (5.5% vs. 2.5%, P = 0.0078), environmental (26.9% vs 16.1%, P <0.001), and rare/other cancers (4.6% vs. 1.9%, P = 0.036) was appreciably higher in NHW compared to NHB participants. When grandparents were included, rates were higher in NHW compared to NHB cases for colorectal (22.8% vs. 16.7%, P = 0.0096), other GI (11.9% vs. 8.0%, P = 0.026), estrogen-driven (29.1% vs 21.2%, P = 0.0018), hematologic (7.5% vs. 2.9%, P = 0.00051), environmental (37.8% vs 20.1%, P <0.001), and rare/other cancers (6.1% vs. 3.1%, P = 0.013). In contrast, the proportion of cases reporting FH of unknown cancer types among FD relatives was higher in NHB compared to NHW participants (16.8% vs. 12.1%, P = 0.049). NHW participants reported a FH of Lynch syndrome-related cancers more frequently than NHB participants when considering FD relatives and grandparents combined (35.2% vs. 26.0%, P <0.001).

**Table 2.**
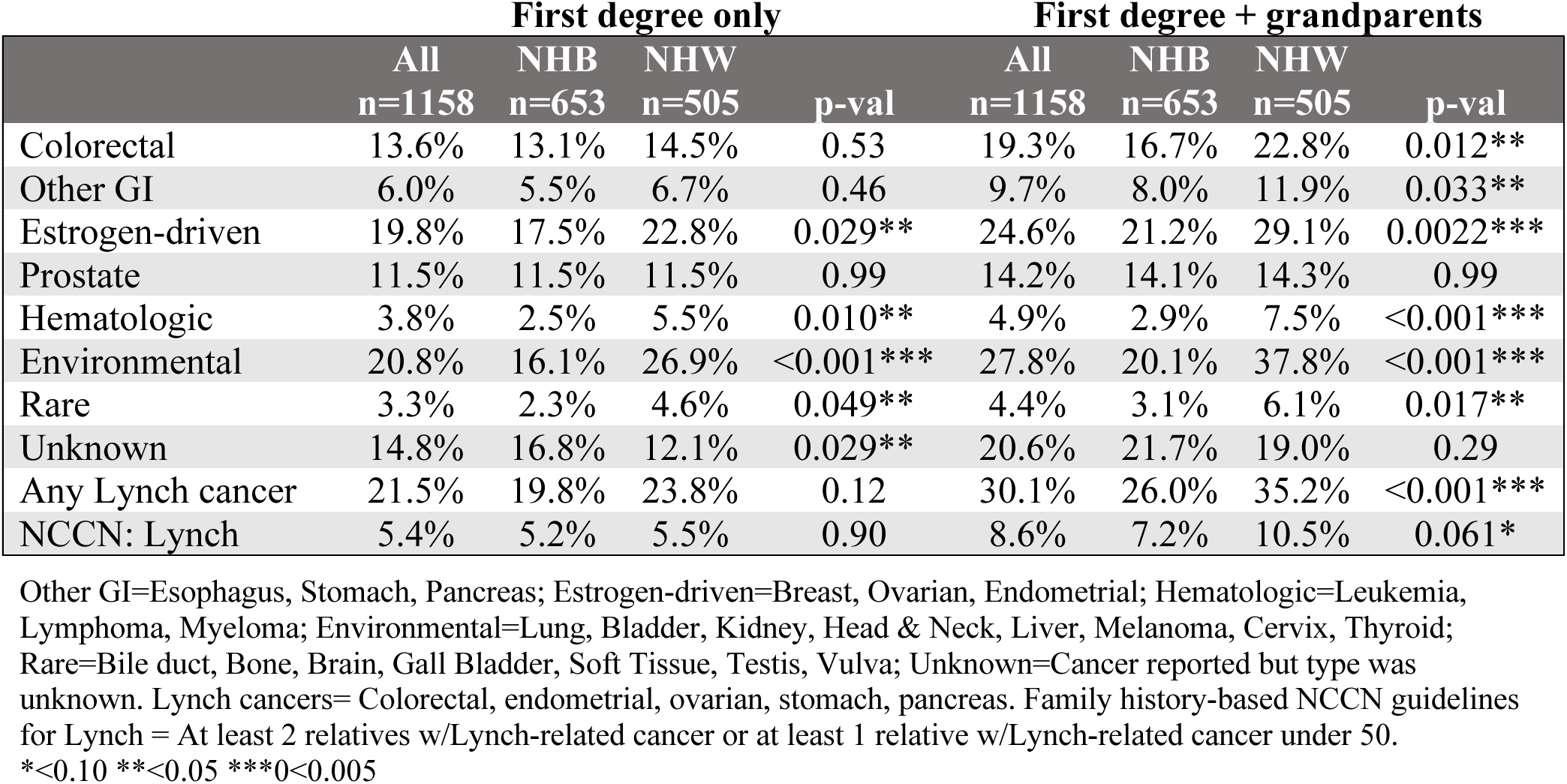
Proportion of cases reporting family history of cancer overall and by race.

We next evaluated cancer FH patterns by age group at diagnosis (Figure 1, eTable 1). In the overall DANCE cohort, the proportion of cases reporting a FD FH of cancer generally increased with age at diagnosis, reaching statistical significance for other GI (P = .04), estrogen-driven (P = .001), environmental (P = .01), and unknown (P = .02) cancers, and for any Lynch syndrome-related cancer (P = .02). The exception was CRC itself: 13.6% of cases diagnosed under 45 reported at least one FD relative with CRC, falling to 9.7% at ages 45-49, and rising again to 12.7% (50-64) and 16.0% (≥65), without reaching statistical significance. Race-stratified analyses showed divergent patterns. In NHB cases, CRC FH was highest at <45 years (22.6%), lowest at 45-49 (8.2%), and intermediate at older ages (11.3% and 14.7%; Fisher exact P = .08). In NHW cases, the pattern was inverted, with CRC FH lowest at <45 (4.0%) and rising with age (11.9%-17.4%; Fisher exact P = .09). Age was also positively associated with FH of estrogen-driven cancers in both groups, with other GI cancers in NHB, and with environmental and unknown cancers in NHW (eTable 1). When FD relatives and grandparents were combined, no significant age-group differences were observed (eTable 1).

**Figure 1.**
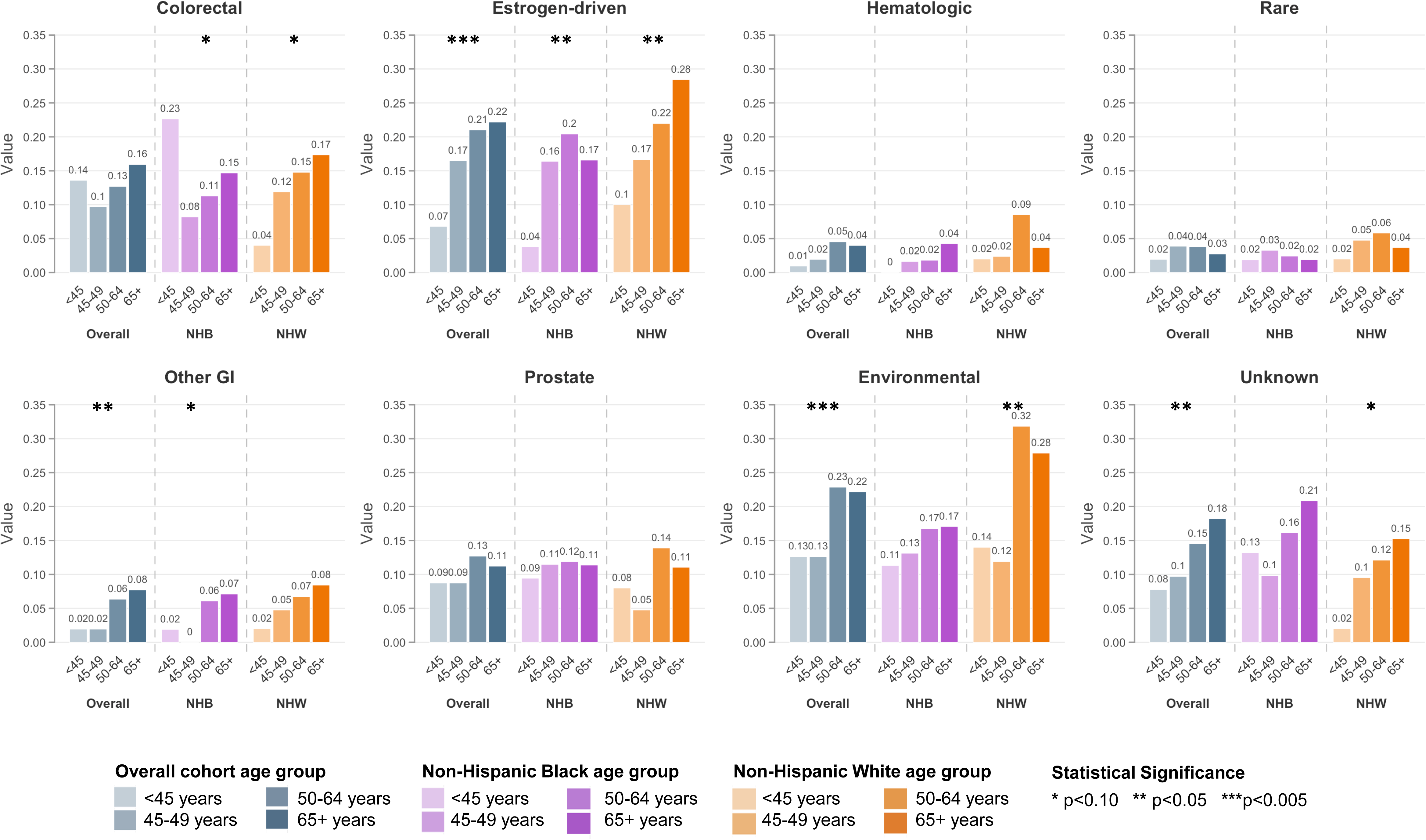
Bar plots indicating the proportion of DANCE cases that reported a first-degree (FD) family history for etiologically-related cancer groupings, overall and stratified by race, are shown. Overall proportions are shown in blue, proportions among NHB are shown in purple, and proportions among NHW are shown in orange. Statistical significance of chi-squared tests of association between site-specific cancer family history and race are indicated by asterisks: *<0.10 **<0.05 ***<0.005. Other GI=Esophagus, Stomach, Pancreas; Estrogen-driven=Breast, Ovarian, Endometrial; Hematologic=Leukemia, Lymphoma, Myeloma; Environmental=Lung, Bladder, Kidney, Head & Neck, Liver, Melanoma, Cervix, Thyroid; Rare=Bile duct, Bone, Brain, Gall Bladder, Soft Tissue, Testis, Vulva; Unknown=Cancer reported but type was unknown.

We then examined associations between FH-based screening criteria and age group at diagnosis (race-stratified, FD relatives only). The three criteria were (1) at least one FD relative with CRC (colon early-screening criterion), (2) any FH of Lynch syndrome-related cancers, and (3) meeting NCCN testing criteria for Lynch syndrome (Table 3). Compared with cases diagnosed at ≥65 years, NHB cases <45 years were marginally more likely to meet the colon early-screening criterion (OR, 1.74; 95% CI, 0.93-3.25; P = .08), while NHW cases showed the opposite pattern (OR, 0.29; 95% CI, 0.11-0.78; P = .01). Among NHB cases, the colon early-screening criterion was less common in those diagnosed at ages 45-49 versus ≥65 (OR, 0.51; 95% CI, 0.21-1.21; P = .13), although this did not reach significance. Any Lynch FH and meeting NCCN testing criteria were similarly distributed across age groups in both racial groups (Table 3), and FD-plus-grandparent definitions did not change these patterns.

**Table 3.**
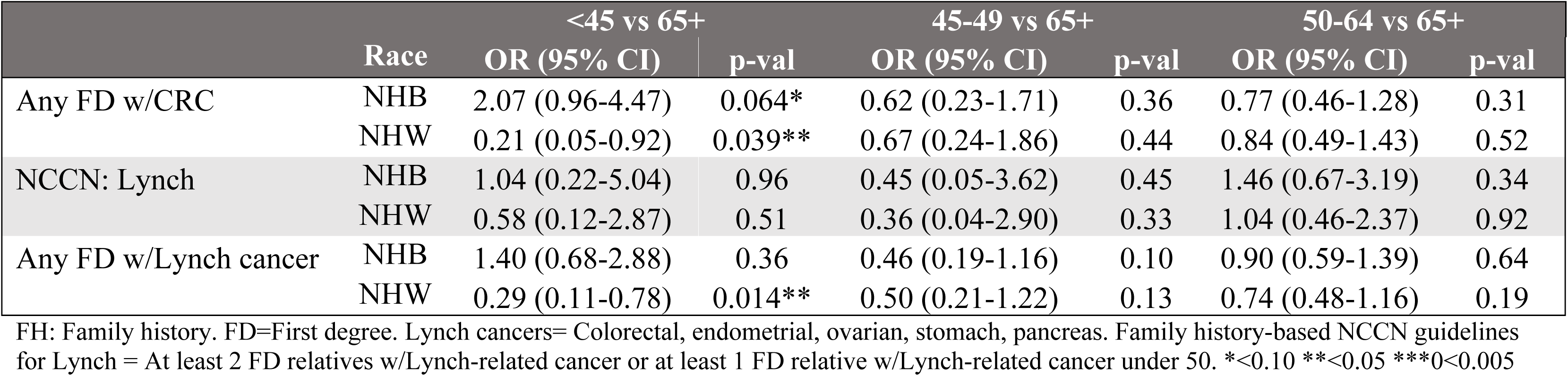
Case-only associations between FH-based screening eligibility criteria and age of onset by race.

|  | Race | <45 vs 65+ |  | 45-49 vs 65+ |  | 50-64 vs 65+ |  |
| --- | --- | --- | --- | --- | --- | --- | --- |
|  |  | OR (95% CI) | p-val | OR (95% CI) | p-val | OR (95% CI) | p-val |
| Any FD w/CRC | NHB | 2.07 (0.96-4.47) | 0.064* | 0.62 (0.23-1.71) | 0.36 | 0.77 (0.46-1.28) | 0.31 |
|  | NHW | 0.21 (0.05-0.92) | 0.039** | 0.67 (0.24-1.86) | 0.44 | 0.84 (0.49-1.43) | 0.52 |
| NCCN: Lynch | NHB | 1.04 (0.22-5.04) | 0.96 | 0.45 (0.05-3.62) | 0.45 | 1.46 (0.67-3.19) | 0.34 |
|  | NHW | 0.58 (0.12-2.87) | 0.51 | 0.36 (0.04-2.90) | 0.33 | 1.04 (0.46-2.37) | 0.92 |
| Any FD w/Lynch cancer | NHB | 1.40 (0.68-2.88) | 0.36 | 0.46 (0.19-1.16) | 0.10 | 0.90 (0.59-1.39) | 0.64 |
|  | NHW | 0.29 (0.11-0.78) | 0.014** | 0.50 (0.21-1.22) | 0.13 | 0.74 (0.48-1.16) | 0.19 |

FH: Family history. FD=First degree. Lynch cancers= Colorectal, endometrial, ovarian, stomach, pancreas. Family history-based NCCN guidelines for Lynch = At least 2 FD relatives w/Lynch-related cancer or at least 1 FD relative w/Lynch-related cancer under 50. \*<0.10 \*\*<0.05 \*\*\*<0.005

Finally, we conducted race- and age-stratified analyses comparing FH-based CRC risk stratification criteria between DANCE CRC cases and 1,434 cancer-free controls from the metropolitan Detroit INHALE study^11^ (**Figure 2**, **eTable 2**). Demographics and overall FD cancer FH patterns for controls are shown in **eTable 3**. Among NHB participants, having at least one FD relative with CRC was associated with higher odds of CRC under the age of 45 (OR=1.44, 95% 1.09-1.89, P = 0.011), at ages 50-64 (OR=1.14, 95% CI 0.,99-1.29, P = 0.053), and at ages 65 and older (OR=1.25, 95% CI 1.06-1.46, P = 0.0070). In contrast, among NHW participants, having at least one FD relative with CRC was only associated with higher odds of CRC for age 45-49 (OR=1.52, 95% CI 1.01-2.28, P = 0.047) and 50-64 (OR=1.14, 95% CI 1.01- 1.28, P = 0.033). Having at least one FD relative with a Lynch syndrome-related cancer was only marginally associated with increased odds of CRC for NHB participants aged <45 (OR=1.24 (95% CI 0.96-1.60, P = 0.099) and for NHW participants age 50-64 (OR=1.08, 95% CI 0.99-1.19, P = 0.091) and ages 65+ (OR=1.11, 95% CI 1.00-1.22, P = 0.052). Meeting NCCN guidelines for Lynch syndrome testing based on FD FH was not significantly associated with odds of CRC.

**Figure 2.**
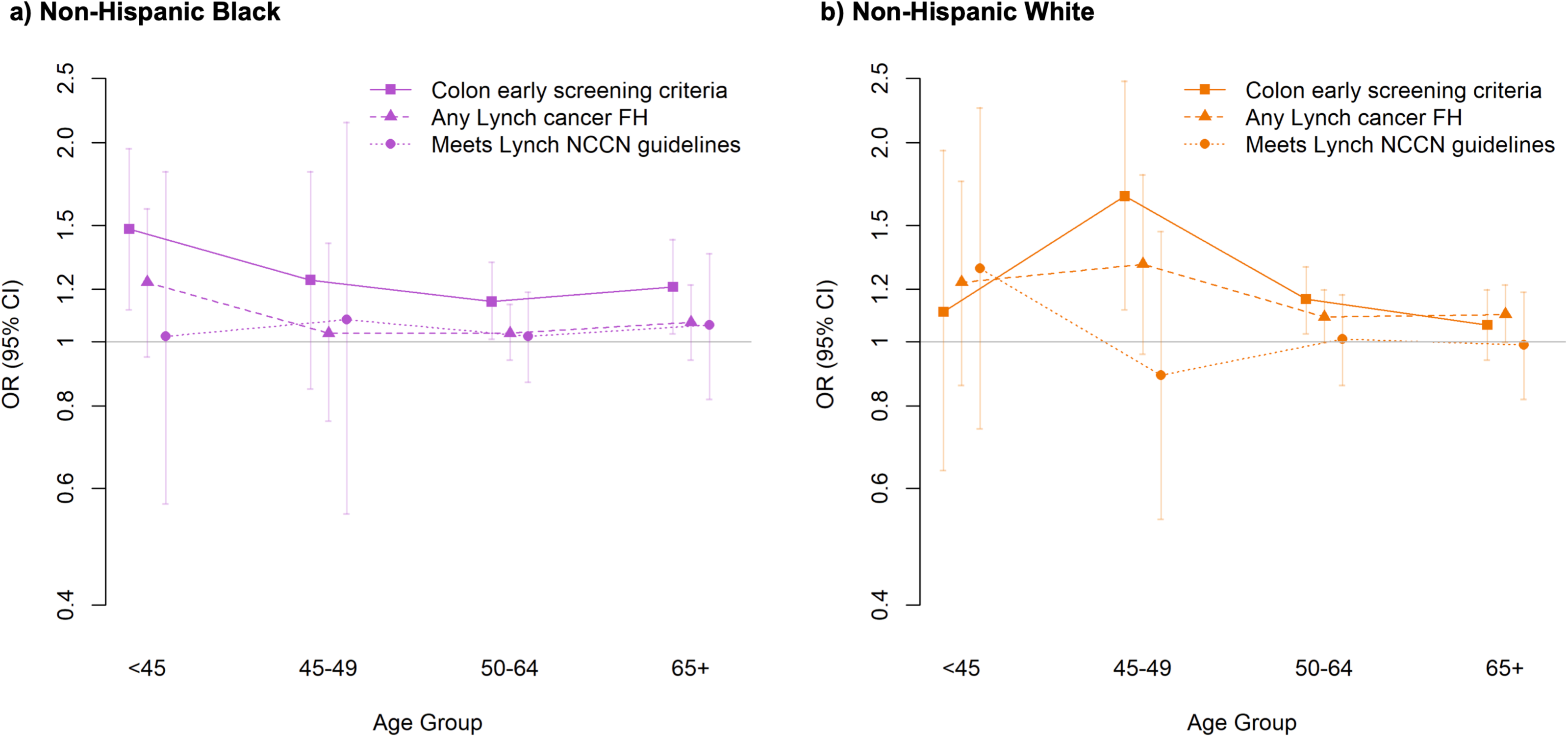
Line plots representing associations between family history (FH)-based screening eligibility criteria and odds of CRC are shown, stratified by age group. Colon early screening criteria = at least one first degree (FD) relative with colorectal cancer (CRC) at any age. Any Lynch cancer FH = at least one FD relative with colorectal, endometrial, ovarian, stomach, or pancreatic cancer at any age. Meets Lynch NCCN guidelines = At least 2 relatives w/Lynch-related cancer or at least 1 relative w/Lynch-related cancer under 50. Odds ratios are representing by squares (colon early screening criteria), triangles (any Lynch cancer FH), and circles (meetings Lynch NCCN guidelines). Vertical lines and dashes represent 95% confidence intervals (CI). Lines connecting age-specific odds ratios are to help visualize trends within screening criteria across age groups. The horizontal gray line represents the null value of 1.0. Panel (a) shows estimates for non-Hispanic Black participants in purple and (b) shows estimates for non-Hispanic white participants in orange.

These data also capture the relative proportions of both cases and controls under age 45 – before screening initiation for average risk individuals – who met clinical criteria for identifying individuals at risk for CRC (**eTable 2**). Specifically, the proportion of cases under 45 with a FD FH of CRC was relatively low overall, although higher among NHB than NHW individuals (22.6% vs 4.0%). In comparison, ∼95% of controls reported no FH of CRC among FD relatives. When criteria were expanded to include any Lynch-related cancer, the proportion of cases under 45 with a FD FH was higher than when considering CRC alone (NHB: 24.5%, NHW: 10.0%); however, the proportion of controls with a FD FH of Lynch-related cancers also rose (11.4% and 7.8%, respectively).

## Discussion

We found that reported cancer FH in individuals with CRC differed substantially by race, that FH burden was generally positively associated with age at diagnosis, and that current FH-based criteria performed poorly in identifying individuals at risk for CRC, particularly for very young onset cases, despite an association with elevated odds of CRC in some groups. To our knowledge, this is one of the first studies to systematically evaluate the sensitivity and specificity of FH-based CRC screening criteria across both age-at-diagnosis and racial groups in a population-based cohort, and our findings highlight important limitations of current approaches that have direct implications for early-onset CRC prevention.

The most clinically actionable finding from this study is that the criterion most commonly used in clinical practice to recommend earlier screening initiation — having at least one FD relative with CRC — failed to identify >80% of very young CRC patients as above average risk. More than three quarters of NHB and nearly all NHW early-onset cases did not report a FD relative with CRC, indicating that the vast majority of individuals who stand to benefit most from earlier screening would not have been identified by this criterion alone. This reflects a fundamental structural limitation of relying on FH as the primary gateway to earlier screening among young people^6,12,13^. The temporal mismatch between the patient’s age at diagnosis and the maturation of their family’s cancer history is an inherent constraint of using FH-based criteria, and it argues for complementary risk stratification approaches that do not depend on FH alone^14,15^.

Expanding criteria to include any Lynch-related cancer in the family captured more very young cases, particularly among NHB individuals, but it also increased the number of controls that would be flagged as high risk. In fact, meeting formal NCCN criteria for Lynch syndrome genetic testing performed least well across CRC risk stratification criteria, capturing under 5% of early-onset cases regardless of race. This suggests that NCCN Lynch testing criteria, while appropriate for their intended purpose of identifying hereditary syndrome carriers, are not well-suited as a population-level tool for flagging individuals who would benefit from earlier CRC screening^16–18^. The two objectives — identifying hereditary syndrome carriers and identifying individuals at sufficient risk to warrant earlier screening — are related but distinct, and criteria optimized for the former may systematically underperform for the latter^19,20^.

A particularly important and novel finding was the divergence in FH patterns between NHB and NHW participants. NHW with CRC diagnosed at older age were more likely than NHB to report FH of CRC, while among young CRC cases, NHB patients were more likely to report a FH compared with young NHW. This is not easily explained by underreporting alone. Black individuals are well documented to report cancer FH less completely than White individuals^21,22^, reflecting lower rates of cancer diagnosis in earlier generations, fragmented family communication around cancer, and structural barriers to genetic services^24,25^. If anything, such underreporting would be expected to attenuate the FH-CRC association we observed in NHB participants, suggesting the true effect may be larger. The biological underpinnings of this divergence remain unclear but may relate to differences in germline variant frequencies and somatic landscapes between ancestry groups^27,28,29^.

Strengths of this study include the population-based ascertainment of CRC cases through cancer registries, systematic collection of cancer FH across a detailed list of cancer sites, and inclusion of sufficient numbers of both NHB and NHW participants to support race-stratified analyses.

Limitations include reliance on self-reported family history, which is known to be incomplete and may be reported less completely by Black than White individuals, potentially attenuating observed associations. The INHALE control group was drawn from Detroit only, so case-control comparisons reflect that catchment area. Finally, FH-based criteria were applied retrospectively; their prospective use in CRC screening decision-making may differ.

In summary, our findings demonstrate that current FH-based criteria for early CRC screening only capture relatively small proportions of CRC cases, particularly among individuals diagnosed before age 45, and that their performance differs significantly by race in ways that are not fully explained by underreporting. These results support a reassessment of how FH criteria are used in CRC risk stratification and point to the need for complementary approaches, including broader population-based screening starting at younger ages and integration of genomic risk information, to reduce the burden of early-onset CRC, particularly in underrepresented populations.

## Data Sharing Statement

De-identified individual participant data underlying the findings reported in this article will be made available to qualified investigators on reasonable request and following execution of a data use agreement consistent with the consent provided by participants and Institutional Review Board requirements. Requests should be directed to the corresponding author. A complete data dictionary will accompany shared data.

## Supporting information

eTables 1-3

## Data Availability

De-identified individual participant data underlying the findings reported in this article will be made available to qualified investigators on reasonable request and following execution of a data use agreement consistent with the consent provided by participants and Institutional Review Board requirements. Requests should be directed to the corresponding author.

