## Supplementary material for "Performance of family history-based colorectal cancer screening criteria by race and age at diagnosis in the Disparities and Cancer Epidemiology (DANCE) study": eTables 1-3

### Supplemental Online Content

#### Contents

**eTable 1.** Proportion of cases reporting family history of cancer by proband age group and race

**eTable 2.** Case-control analysis of family history-based CRC screening criteria, by race and age group

**eTable 3.** Demographic characteristics of INHALE control participants by age group

#### eTable 1. Proportion of cases reporting family history of cancer by proband age group and race

| **a) Etiologic groupings** | | | | | | | | | | | | | | | | | | |
|---|---|---|---|---|---|---|---|---|---|---|---|---|---|---|---|---|---|---|
| | **All** | | | | | | **NHB** | | | | | | **NHW** | | | | | |
| | | **<45** | **45-49** | **50-64** | **65+** | | | **<45** | **45-49** | **50-64** | **65+** | | | **<45** | **45-49** | **50-64** | **65+** | |
| **Relatives' cancer site** | | **n=103** | **n=103** | **n=551** | **n=401** | **p-val** | | **n=53** | **n=61** | **n=328** | **n=211** | **p-val** | | **n=50** | **n=42** | **n=223** | **n=190** | **p-val** |
| Colorectal | | 0.214 | 0.204 | 0.189 | 0.192 | 0.918 | | 0.245 | 0.148 | 0.162 | 0.161 | 0.458 | | 0.18 | 0.286 | 0.229 | 0.226 | 0.697 |
| Other GI | | 0.058 | 0.087 | 0.103 | 0.1 | 0.572 | | 0.075 | 0.049 | 0.088 | 0.076 | 0.829 | | 0.04 | 0.143 | 0.126 | 0.126 | 0.298 |
| Estrogen-driven | | 0.184 | 0.272 | 0.247 | 0.254 | 0.45 | | 0.132 | 0.262 | 0.226 | 0.194 | 0.293 | | 0.24 | 0.286 | 0.278 | 0.321 | 0.665 |
| Prostate | | 0.155 | 0.136 | 0.151 | 0.127 | 0.734 | | 0.151 | 0.18 | 0.137 | 0.133 | 0.768 | | 0.16 | 0.071 | 0.17 | 0.121 | 0.277 |
| Hematologic | | 0.068 | 0.029 | 0.053 | 0.045 | 0.599 | | 0.038 | 0.016 | 0.021 | 0.043 | 0.428 | | 0.1 | 0.048 | 0.099 | 0.047 | 0.188 |
| Environmental | | 0.252 | 0.282 | 0.292 | 0.264 | 0.747 | | 0.17 | 0.23 | 0.213 | 0.18 | 0.684 | | 0.34 | 0.357 | 0.408 | 0.358 | 0.68 |
| Rare | | 0.039 | 0.049 | 0.054 | 0.03 | 0.324 | | 0.019 | 0.033 | 0.037 | 0.024 | 0.836 | | 0.06 | 0.071 | 0.081 | 0.037 | 0.283 |
| Unknown | | 0.146 | 0.175 | 0.205 | 0.229 | 0.243 | | 0.17 | 0.18 | 0.223 | 0.232 | 0.72 | | 0.12 | 0.167 | 0.179 | 0.226 | 0.34 |
| b) Syndrome-related groupings | | | | | | | | | | | | | | | | | | |
| Any Lynch cancer: FD+GP | | 0.301 | 0.301 | 0.296 | 0.307 | 0.986 | | 0.321 | 0.23 | 0.268 | 0.242 | 0.618 | | 0.28 | 0.405 | 0.336 | 0.379 | 0.479 |
| NCCN guidelines for Lynch w/FD+GP | | 0.058 | 0.068 | 0.093 | 0.08 | 0.681 | | 0.057 | 0.016 | 0.085 | 0.066 | 0.267 | | 0.06 | 0.143 | 0.103 | 0.095 | 0.606 |

Other GI=Esophagus, Stomach, Pancreas; Estrogen-driven=Breast, Ovarian, Endometrial; Hematologic=Leukemia, Lymphoma, Myeloma; Environmental=Lung, Bladder, Kidney, Head & Neck, Liver, Melanoma, Cervix, Thyroid; Rare=Bile duct, Bone, Brain, Gall Bladder, Soft Tissue, Testis, Vulva; Unknown=Cancer reported but type was unknown

#### eTable 2. Case-control analysis of family history-based CRC screening criteria, by race and age group

| **a) NHB** | **<45** | | | | **45-49** | | | | **50-64** | | | | **65+** | | | |
|---|---|---|---|---|---|---|---|---|---|---|---|---|---|---|---|---|
| | **Cases** | **Controls** | | | **Cases** | **Controls** | | | **Cases** | **Controls** | | | **Cases** | **Controls** | | |
| | **n (%)** | **n (%)** | **OR (95% CI)** | **p-val** | **n (%)** | **n (%)** | **OR (95% CI)** | **p-val** | **n (%)** | **n (%)** | **OR (95% CI)** | **p-val** | **n (%)** | **n (%)** | **OR (95% CI)** | **p-val** |
| **Any FD w/CRC** | | | | | | | | | | | | | | | | |
| Yes | 12 (22.6) | 2 (4.5) | Crude: 1.44 (1.09-1.89) | 0.011 | 5 (8.2) | 2 (4.2) | Crude: 1.18 (0.80-1.73) | 0.4 | 37 (11.3) | 26 (7.1) | Crude: 1.14 (0.99-1.29) | 0.053 | 31 (14.7) | 10 (6.0) | Crude: 1.25 (1.06-1.46) | 0.007 |
| No | 41 (77.3) | 42 (95.5) | Adjusted: 1.48 (1.12-1.96) | 0.008 | 56 (91.8) | 46 (95.8) | Adjusted: 1.24 (0.85-1.81) | 0.26 | 291 (88.7) | 342 (92.9) | Adjusted: 1.15 (1.01-1.32) | 0.034 | 180 (85.3) | 157 (94.0) | Adjusted: 1.21 (1.03-1.43) | 0.019 |
| NCCN guidelines for Lynch | | | | | | | | | | | | | | | | |
| Yes | 2 (3.8) | 1 (2.3) | Crude: 1.13 (0.64-2.02) | 0.67 | 1 (1.6) | 1 (2.1) | Crude: 0.94 (0.47-1.90) | 0.87 | 21 (6.4) | 21 (5.7) | Crude: 1.03 (0.88-1.21) | 0.7 | 10 (4.7) | 6 (3.6) | Crude: 1.07 (0.84-1.38) | 0.58 |
| No | 51 (96.2) | 43 (97.7) | Adjusted: 1.02 (0.57-1.81) | 0.95 | 60 (98.4) | 47 (97.9) | Adjusted: 1.08 (0.55-2.15) | 0.82 | 307 (93.6) | 347 (94.3) | Adjusted: 1.02 (0.87-1.19) | 0.84 | 201 (95.3) | 161 (96.4) | Adjusted: 1.06 (0.82-1.36) | 0.66 |
| Any FD w/Lynch cancer | | | | | | | | | | | | | | | | |
| Yes | 13 (24.5) | 5 (11.4) | Crude: 1.24 (0.96-1.60) | 0.099 | 6 (9.8) | 5 (10.4) | Crude: 0.98 (0.72-1.35) | 0.92 | 64 (19.5) | 62 (16.8) | Crude: 1.05 (0.95-1.15) | 0.36 | 46 (21.8) | 27 (16.2) | Crude: 1.09 (0.96-1.24) | 0.17 |
| No | 40 (75.4) | 39 (88.6) | Adjusted: 1.23 (0.95-1.59) | 0.13 | 55 (90.2) | 43 (89.6) | Adjusted: 1.03 (0.76-1.41) | 0.83 | 264 (80.5) | 306 (83.2) | Adjusted: 1.03 (0.94-1.14) | 0.51 | 165 (78.2) | 140 (83.8) | Adjusted: 1.07 (0.94-1.22) | 0.3 |
| b) NHW | <45 | | | | 45-49 | | | | 50-64 | | | | 65+ | | | |
| | Cases | Controls | | | Cases | Controls | | | Cases | Controls | | | Cases | Controls | | |
| | n (%) | n (%) | OR (95% CI) | p-val | n (%) | n (%) | OR (95% CI) | p-val | n (%) | n (%) | OR (95% CI) | p-val | n (%) | n (%) | OR (95% CI) | p-val |
| Any FD w/CRC | | | | | | | | | | | | | | | | |
| Yes | 2 (4.0) | 2 (3.9) | Crude: 1.01 | 0.98 | 5 (11.9) | 1 (1.9) | Crude: 1.52 | 0.047 | 33 (14.8) | 38 (9.2) | Crude: 1.14 | 0.033 | 33 (17.4) | 40 (13.7) | Crude: 1.07 | 0.28 |

| | | | | | | | | | | | | | | | | |
|---|---|---|---|---|---|---|---|---|---|---|---|---|---|---|---|---|
| | | | (0.61-1.67) | | | | (1.01-2.28) | | | | (1.01-1.28) | | | | (0.95-1.21) | |
| No | 48 (96.0) | 49 (96.1) | Adjusted: 1.11 (0.64-1.95) | 0.7 | 37 (88.1) | 52 (98.1) | Adjusted: 1.66 (1.12-2.48) | 0.014 | 190 (85.2) | 374 (90.8) | Adjusted: 1.16 (1.03-1.30) | 0.015 | 157 (82.6) | 251 (86.3) | Adjusted: 1.06 (0.94-1.20) | 0.34 |
| NCCN guidelines for Lynch | | | | | | | | | | | | | | | | |
| Yes | 2 (4.0) | 1 (2.0) | Crude: 1.19 (0.67-2.13) | 0.55 | 1 (2.4) | 3 (5.7) | Crude: 0.82 (0.50-1.35) | 0.43 | 14 (6.3) | 26 (6.3) | Crude: 1.00 (0.86-1.16) | 0.99 | 11 (5.8) | 17 (5.8) | Crude: 1.00 (0.83-1.20) | 0.98 |
| No | 48 (96.0) | 50 (98.0) | Adjusted: 1.29 (0.74-2.26) | 0.37 | 41 (97.6) | 50 (94.3) | Adjusted: 0.89 (0.54-1.47) | 0.65 | 206 (93.7) | 386 (93.7) | Adjusted: 1.01 (0.86-1.18) | 0.92 | 179 (94.2) | 274 (94.2) | Adjusted: 0.99 (0.82-1.19) | 0.88 |
| Any FD w/Lynch cancer | | | | | | | | | | | | | | | | |
| Yes | 5 (10.0) | 4 (7.8) | Crude: 1.07 (0.76-1.51) | 0.71 | 7 (16.7) | 4 (7.5) | Crude: 1.25 (0.91-1.70) | 0.17 | 52 (23.3) | 73 (17.7) | Crude: 1.08 (0.99-1.19) | 0.091 | 56 (29.5) | 63 (21.6) | Crude: 1.11 (1.00-1.22) | 0.052 |
| No | 45 (90.0) | 47 (92.2) | Adjusted: 1.23 (0.86-1.75) | 0.25 | 35 (83.3) | 49 (92.5) | Adjusted: 1.31 (0.96-1.79) | 0.096 | 171 (76.7) | 339 (82.3) | Adjusted: 1.09 (1.00-1.20) | 0.059 | 134 (70.5) | 228 (78.4) | Adjusted: 1.10 (1.00-1.22) | 0.057 |
| | Adjusted for numberof first degree relatives and education | | | | | | | | | | | | | | | |

#### eTable 3. Demographic characteristics of INHALE control participants by age group

| | <45 | 45-49 | 50-64 | 65+ | p-val |
|---|---|---|---|---|---|
| **Demographics** | **n (%)** | **n (%)** | **n (%)** | **n (%)** | |
| **Race** | | | | | |
| Non-Hispanic Black | 44 (46.3) | 48 (47.5) | 368 (47.2) | 167 (36.5) | 0.002 |
| Non-Hispanic White | 51 (53.7) | 53 (52.5) | 412 (52.8) | 291 (63.5) | |
| Sex | | | | | |
| Female | 59 (62.1) | 64 (36.6) | 426 (54.6) | 244 (53.3) | 0.15 |
| Male | 36 (37.9) | 37 (63.4) | 354 (45.4) | 214 (46.7) | |
| Highest level of education | | | | | |
| Less than HS | 11 (11.6) | 11 (10.9) | 71 (9.1) | 28 (6.1) | 0.4 |
| HS | 23 (24.2) | 29 (28.7) | 200 (25.6) | 120 (26.2) | |
| Some college | 51 (53.7) | 50 (49.5) | 416 (53.3) | 236 (51.5) | |
| Professional/graduate | 10 (10.5) | 11 (10.9) | 93 (11.9) | 73 (15.9) | |
| Unknown | 0 (0) | 0 (0) | 0 (0) | 1 (0.2) | |
| No. of first degree relatives | | | | | |
| Median (Min-Max) | 5 (1-16) | 6 (2-19) | 7 (1-23) | 7 (2-20) | <0.001 |
| Family history* | | | | | |
| Colorectal | 0.042 | 0.03 | 0.082 | 0.109 | 0.019 |
| Other GI | 0.032 | 0.05 | 0.078 | 0.081 | 0.29 |
| Estrogen-driven | 0.084 | 0.129 | 0.192 | 0.188 | 0.026 |
| Prostate | 0.074 | 0.109 | 0.09 | 0.094 | 0.85 |
| Hematologic | 0.011 | 0.02 | 0.04 | 0.059 | 0.096 |
| Environmental | 0.147 | 0.198 | 0.251 | 0.221 | 0.092 |
| Rare | 0.042 | 0.04 | 0.044 | 0.061 | 0.57 |
| Unknown | 0.021 | 0.04 | 0.094 | 0.12 | 0.002 |
| Any Lynch cancer | 0.095 | 0.089 | 0.173 | 0.197 | 0.01 |
| NCCN guidelines for Lynch | 0.021 | 0.04 | 0.06 | 0.05 | 0.43 |

* Proportion of cases reporting family history of cancer type among FD relatives
